# COVID-19 Vaccine Hesitancy Among Patients with Inflammatory Bowel Disease on Biologic Therapies; A cross-sectional study

**DOI:** 10.1101/2021.12.03.21267237

**Authors:** Mohammad Shehab, Yasmin Zurba, Ali Al Abdulsalam, Ahmad Alfadhli, Sara Elouali

## Abstract

**Background:** COVID-19 Vaccinations have been shown to be effective in reducing risk of severe infection, hospitalization, and death. They also have been shown to be safe and effective in patients with inflammatory bowel disease (IBD) on biologic therapies. In this study, we aimed to evaluate the prevalence of vaccination among patients with IBD on biologic therapies.

**Methods:** A single-center prospective cross-sectional study conducted at a tertiary care inflammatory bowel disease center. Data from patients with inflammatory bowel disease (IBD) who attended the gastroenterology infusion clinic from June 1st, 2021 until October 31st, 2021 were retrieved. Patients received infliximab or vedolizumab at least 6 weeks before recruitment were included. The primary outcome was prevalence of COVID-19 vaccination. The secondary outcome was to assess whether prevalence of COVID-19 vaccination differed based sex, age, type of biologic therapy and citizenship status.

**Results:** The total number of inflammatory bowel disease (IBD) patients enrolled in the study was 280 (56.0% male and 44.0% female). The median age was 33.2 years and BMI was 24.8 kg/m^2^. 112 patients with ulcerative colitis (40.0%) and 168 (60.0%) with Crohn’s disease. 117 (41.8%) were vaccinated with either BNT162b2 or ChAdOx1 nCoV-19 and 163 (58.2%) were not vaccinated. Female patients were more likely to receive the vaccine compared to male patients (83.0% vs. 63.8%, p < 0.001). In addition, older patients (above the age 50) were also more likely to receive the vaccine than younger patients, below the age of 50 (95.6% vs 31.2% p< 0.001). Expatriates were more likely to receive the vaccine than citizens (84.8% vs 25.0%, p < 0.001). There was no statistical difference between patients on Infliximab and vedolizumab in terms of prevalence of vaccination (40.0% vs 48.0%, p= 0.34).

**Conclusion:** The overall prevalence of COVID-19 vaccination among patients with inflammatory bowel disease (IBD) on biologic therapies was lower than the general population and world health organization (WHO) recommendation. Female patients, patients above the age of 50, and expatriates were more likely to be vaccinated. On the other hand, male patients, patients below the age of 50, and citizens were less likely to be vaccinated.

## 1. Introduction

The global spread of Coronavirus disease 2019 (COVID-19) has impacted every aspect of life. It was declared a pandemic by the World Health Organization (WHO) on the 11th of March, 2020 (1). Although the pandemic has not finished yet, the most effective way of ending it is with widespread vaccination (2). Widespread coverage of vaccines may be hindered by vaccine hesitancy, defined by WHO as a “delay in acceptance or refusal of vaccination despite availability of vaccination services.”(3). Patients who suffer from immune-mediated inflammatory diseases, such as the inflammatory bowel diseases (IBD), are often treated with biologic therapies. Those patients may be hesitant to receive COVID-19 vaccines due to safety and efficacy concerns (4).

Interestingly, it has been shown in a recent meta-analysis that biologic therapies are not associated with severe COVID-19 or worse outcomes in patients with IBD (5). In addition, COVID-19 vaccines have been shown to be effective in patients with IBD on biologics (6,7). The Surveillance Epidemiology of Coronavirus Under Research Exclusion (SECURE-IBD) is an international database that was established at the beginning of the COVID-19 pandemic to monitor and report outcomes of COVID-19 infection in IBD patients. This registry includes the outcomes of more than 6000 IBD patients infected with COVID-19 from 72 countries (8). The SECURE-IBD study showed that in patients with IBD and COVID-19 infection, biologic therapies are not associated with poor outcome (9). The International Organization for the study of Inflammatory Bowel Disease (IOIBD) recommends COVID-19 vaccine for patients with IBD (10).

In Kuwait, BNT162b2 (Pfizer/BioNTech) and ChAdOx1 nCoV-19 (AstraZeneca) vaccines are the only two approved COVID-19 vaccines since December 2020. The eligibility to receive BNT162b2 vaccine is 12 years and older, whereas for ChAdOx1 nCoV-19 is 18 years and older. By the end of October 2021, the government of Kuwait announced that 74.0% of the eligible population in Kuwait had received at least one dose of either COVID-19 vaccine(11). However, there is limited information on COVID-19 vaccine prevalence in patients with IBD who are receiving biologic therapies. The aim of this study is to evaluate the prevalence of COVID-19 vaccines in patients with IBD on biologic therapies and evaluate possible factors that are associated with vaccination.

## 2. Material and Methods

### Study design and recruitment

A single-center prospective cross-sectional study conducted at Mubarak Al-Kaber University Hospital, a tertiary care inflammatory bowel disease center. Data from patients with inflammatory bowel disease (IBD) who attended the gastroenterology infusion clinic from June 1st, 2021 until October 31st, 2021 were retrieved. Patients were eligible to be included if they: 1) had a confirmed diagnosis of inflammatory bowel disease before the start of the study 2) received infliximab or vedolizumab at least 6 weeks before recruitment 3) are 18 years of age or older. Exclusion criteria were the following: 1) prior suspected or confirmed severe acute respiratory syndrome Coronavirus 2 (SARS-CoV-2) infection 2) corticosteroid use within two weeks of recruitment date 3) contraindication to receiving the COVID-19 vaccine as per the Food and Drug Administration (FDA) recommendations (12).

The study was performed and reported in accordance with Strengthening the Reporting of Observational Studies in Epidemiology (STROBE) guidelines (supplementary 1). Patient and disease characteristics, demographics, and vaccination details were obtained from electronic medical records. Diagnosis of IBD was made according to the international classification of diseases (ICD-10 version:2019). Patients were considered to have IBD when they had ICD-10 K50, K50.1, K50.8, K50.9 corresponding to Crohn’s disease (CD) and ICD-10 K51, K51.0, K51.2, K51.3, K51.5, K51.8, K51.9 corresponding to ulcerative colitis (UC)(13).

The primary outcome was the prevalence of COVID-19 vaccination among patients with IBD receiving infliximab or vedolizumab. The secondary outcome was to assess whether prevalence of COVID-19 vaccination differed based sex, age, type of biologic therapy and citizenship status.

Analyses were conducted using R (R core team, 2017). The statistical significance level was set at α = 0.05. Descriptive analyses were conducted to calculate frequencies and proportions of categorical variables. χ2 tests were used to assess whether prevalence of COVID-19 vaccination differed across categories of demographic variables.

## 3. Results

The total number of inflammatory bowel disease (IBD) patients enrolled in the study was 280 (56.0% male and 44.0% female). Median age was 33.2 years, median BMI was 24.8 kg/m^2^, and 58 (20.0%) of the participants were smokers. The most common comorbidities were asthma (13.6%), diabetes (6.7%), and arthritis (5.0%). The median duration of therapy at the time of the study was 12 months for Infliximab and 11 months for Vedolizumab. The participants included: 112 patients with ulcerative colitis (UC) (40.0%) and 168 (60.0%) with Crohn’s disease (CD) (Table 1).

**Table 1:** Demographics of Patients with IBD

| **Variable** | **Study Group\* (n=280)** |
| --- | --- |
| Mean age (years) | 33.2 |
| Sex n (%) |  |
| Male | 157 (56.0%) |
| Female | 123 (44.0%) |
| BMI (Median) | 24.8 |
| Smoking n (%) | 58 (20.0%) |
| Comorbidities n (%) |  |
| Diabetes | 19 (6.7%) |
| OSA | 5 (1.7%) |
| Hypertension | 9 (3.2%) |
| Cardiovascular Disease | 9 (3.2%) |
| Arthritis | 14 (5.0%) |
| Kidney | 9 (3.2%) |
| Asthma | 38 (13.6%) |
| Hyperlipidemia | 9 (3.2%) |
| Median infliximab therapy (months) | 12 |
| Median vedolizumab therapy (months) | 11 |
| Disease extent, n (%) |  |
| Ulcerative colitis (UC) | 112 (40.0%) |
| E1: ulcerative proctitis | 20 (17.8%) |
| E2: left sided colitis | 32 (28.5%) |
| E3: extensive colitis | 60 (53.6%) |
| Crohn's disease (CD) | 168 (60.0%) |
| L1: ileal | 84 (50.0%) |
| L2: colonic | 20 (11.9%) |
| L3: ileocolonic | 59 (35.2%) |
| L4: upper gastrointestinal | 5 (2.8%) |
| B1: inflammatory | 75 (44.6%) |
| B2: stricturing | 44 (26.2%) |
| B3: penetrating | 49(29.2%) |

Among the participants, 117 (41.8%) were vaccinated with either BNT162b2 or ChAdOx1 nCoV-19 and 163 (58.2%) were not vaccinated. Of the vaccinated patients, 25 (21.4%) were vaccinated with one dose, whereas 92 (78.6%) were vaccinated with two doses. Of the total number of participants, 232 (82.9%) were receiving infliximab, and 48 (17.1%) were receiving vedolizumab. Ninety-four (40.0%) patients on infliximab were vaccinated, whereas 138 (60.0%) were not. On the other hand, 23 (48.0%) patients on vedolizumab were vaccinated and 25 (52.0%) were not. Regarding age, 46 (16.4%) participants were above the age of 50 (44 (95.6%) vaccinated, and 2 (4.4%) unvaccinated), whereas 234 (83.6%) were below the age of 50 (73 (31.2%) vaccinated, and 161 (68.8%) unvaccinated). Regarding nationality, 50 (25.0%) citizens were vaccinated and 151 (75.0%) were not. Among expatriates, 67 (84.8%) were vaccinated and 12 (15.2%) were not. Of the total number of male participants, 98 (62.4%) were vaccinated and 59 (37.6%) were not, and of the female participants, 102 (83.0%) were vaccinated and 21 (17.1%) were not. A total of five patients were pregnant, all of whom were not vaccinated (Table 2).

**Table 2:** Demographics of patients with IBD according to vaccination status

| Demographics/ | Total number of | Vaccinated patients | Unvaccinated patients |
| --- | --- | --- | --- |

| vaccination status | patients (280) | (117) | (163) |
| --- | --- | --- | --- |
| One dose | 25 | 25 | N/A |
| Two doses | 92 | 92 | N/A |
| Infliximab | 232 | 94 (40.0%) | 138 (60.0%) |
| Vedolizumab | 48 | 23 (48.0%) | 25 (52.0%) |
| Age >50 | 46 | 44 (95.6%) | 2 (4.4%) |
| Age<50 | 234 | 73 (31.2%) | 161 (68.8%) |
| Citizens | 201 | 50 (25.0%) | 151 (75.0%) |
| Expatriates | 79 | 67 (84.8%) | 12 (15.2%) |
| Male | 157 | 98 (62.4%) | 59 (37.6%) |
| Female | 123 | 102 (83.0%) | 21 (17.1%) |
| Pregnant | 5 | 0 | 5 |

Female patients were more likely to receive the vaccine compared to male patients (83.0% vs. 63.8%, p< 0.001). In addition, participants of older age (above the age of 50) were also more likely to receive the vaccine than younger patients, below the age of 50 years old (95.6% vs 31.2% p< 0.001). Expatriates were more likely to receive the vaccine than citizens (84.8% vs 25.0%, p < 0.001) (Table 2, Figure 1). However, there was no statistical difference between patients on Infliximab and vedolizumab in terms of prevalence of vaccination (40.0% vs 48.0%, p= 0.34) (Table 2, figure 2).

**Figure 1:**
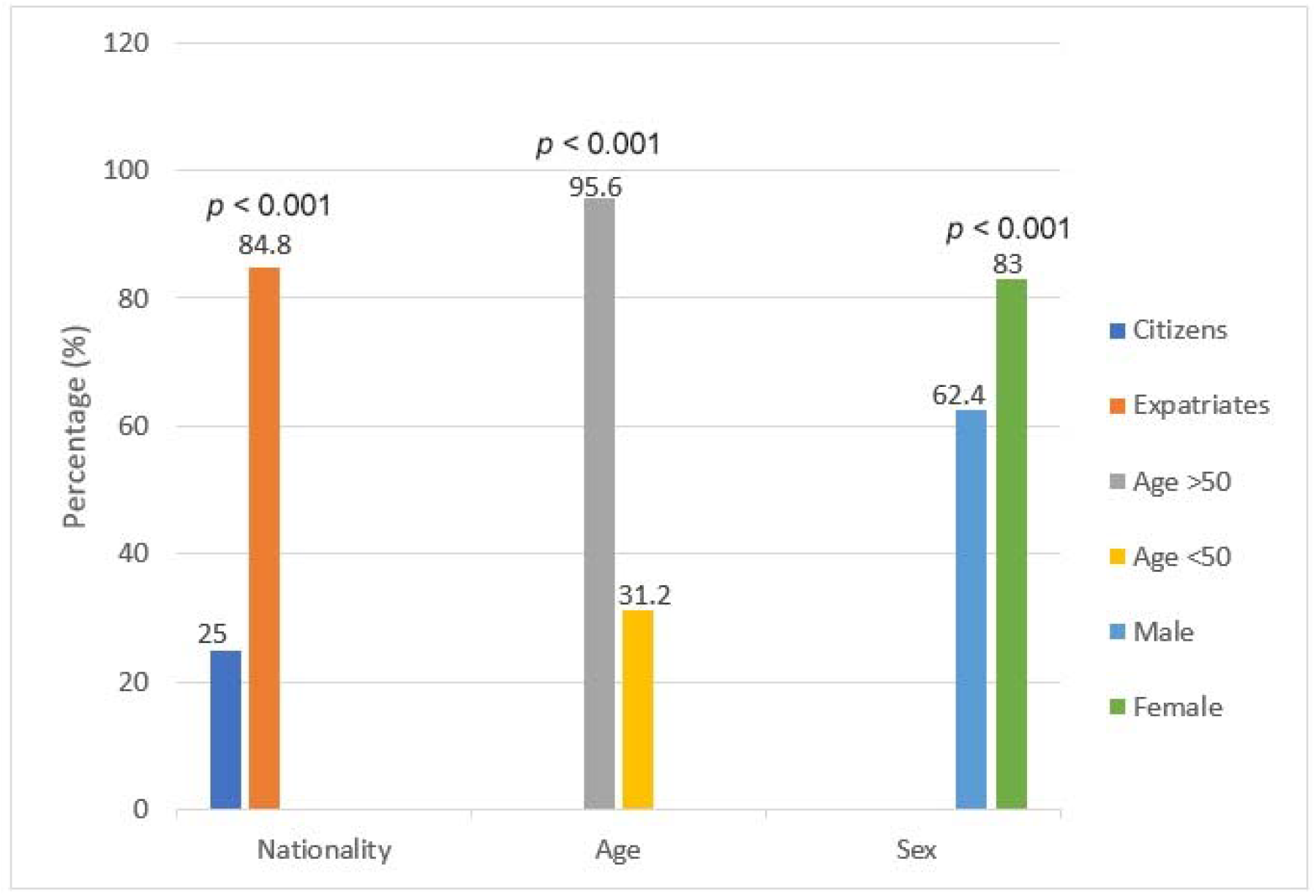
Demographics of patients with IBD based on vaccination status.

**Figure 2:**
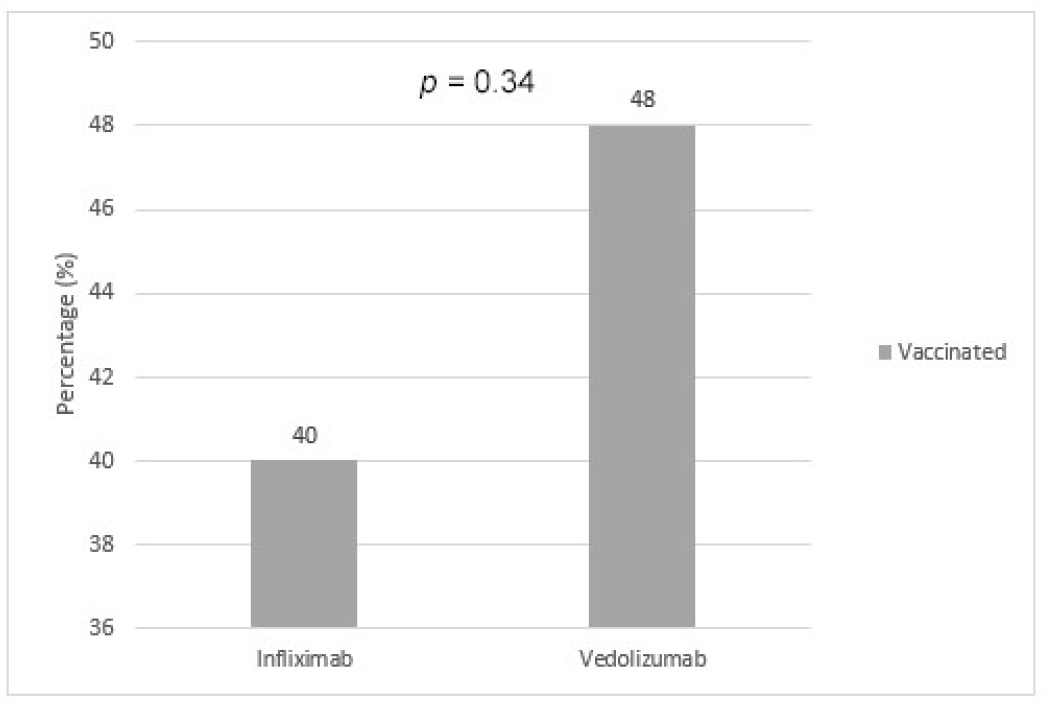
Vaccination status of patients with IBD based on type of biologic therapy.

## 4. Discussion

The world’s leading inflammatory bowel disease (IBD) medical organizations recommend COVID-19 vaccination among IBD patients on biologic therapies; however, for unclear reasons, there remains an evident hesitancy to receiving the vaccine (14).

In this study, we report that among 280 IBD patients on intravenous biologic therapy, 117 (41.8%) were vaccinated and 163 (58.2%) were not vaccinated. According to World Health Organization (WHO) experts, the target vaccination rate in the population of every country is 60-80% in order to achieve herd immunity and break the chain of transmission(15,16). A cross-sectional study conducted in Saudi Arabia in October 2021 reported that the majority of IBD patients (68.0%) had received the COVID-19 vaccine, which is higher than the uptake found in our study (17).

In our study, positive predictors of willingness to receive COVID-19 vaccination were found to be female sex, older age (above 50 years old), and expatriates. Negative predictors were male sex, younger age (below 50 years old), and citizens. Females were more willing to receive the COVID-19 vaccine in relation to males. This could be explained by the fact that females were found to have higher rates of medical services utilization, including healthcare services for prevention, detection, and treatment (18). In addition, females make up the majority of frontline healthcare workers, putting them at higher occupational exposure (19). A possible rationale for higher vaccination rate among patients above the age of 50 is the higher prevalence of comorbidities in this age group and fear of severe COVID-19 infection. This is consistent with the available data in the literature, as older age has been associated with higher COVID-19 vaccination rates (20).The vaccination rate among expatriate participants was higher than that of citizens. This may be due to the fact that expatriate participants are more likely to travel regularly to their home countries to visit their families and local health regulations allow only vaccinated individuals to travel abroad. Moreover, most expatriates work in the private sector, which requires positive vaccination status to return to work whereas citizens mainly work at the government, where vaccination is not mandatory yet. In our study, we noticed that the percentage of vaccinated patients receiving Infliximab and Vedolizumab were similar, 40.0% and 48.0% respectively. A cohort study conducted in the United Kingdom (UK) also showed similar vaccine uptake among IBD patients treated with infliximab and vedolizumab (21). A German case-control study distributed a questionnaire that explores willingness or hesitancy to receive any COVID-19 vaccine to two groups: a group of patients with IBD and a group of healthcare-affiliated volunteers without IBD. The participants in the IBD group were significantly more hesitant towards receiving the COVID-19 vaccine (58.5%) compared to the control group (65.1%) (22).

Females of child-bearing age or in the peripartum period were found to have lower vaccination rates (21) which is in line with our study’s findings in which none of the pregnant participants were vaccinated. This may be due to fear of long-term effects on the mother or fetus despite that the vaccines were recommended by the Royal College of Obstetricians and Gynecologists (RCOG) and the American College of Obstetricians and Gynecologists (ACOG) (23,24). A cohort study was conducted at St. George’s University Hospitals National Health Service Foundation Trust, London, United Kingdom where 1328 pregnant females were followed up, of whom only 140 females agreed to receive the COVID-19 vaccine. It was evident that 10.5% of participants agreed to receive the vaccine, leaving 89.5% hesitant. The results of this study support the evidence that receiving COVID-19 vaccination in pregnancy does not change perinatal outcomes (25). A multimethods study on females’ acceptance towards COVID-19 vaccination resulted in 81.2% acceptance rate among non-pregnant females. This number drops significantly to 62.1% in pregnant females and was attributed to vaccine safety concerns (26). The literature is lacking with regards to administering COVID-19 vaccination to lactating females with IBD since lactating females were excluded from initial clinical trials. For this reason, there is insufficient evidence on the effects of vaccination on lactating females and newborns. The COVID-19 vaccines are non-live vaccines and are therefore expected to be safe in lactating females and newborns (27,28). Since breastfeeding has many benefits, it is recommended by ACOG, Joint Committee on Vaccination and Immunization (JCVI), Centers for Disease Control and Prevention (CDC), Society of Obstetricians and Gynaecologists of Canada (SOGC) to initiate or resume breastfeeding in lactating females who are vaccinated (29).

The International Organization for the study of Inflammatory Bowel Disease (IOIBD) is an organization of clinical researchers devoted to the study and management of IBD (30). An international consensus meeting among members of IOIBD took place to establish reccommendations concerning the use of COVID-19 vaccines in patients with IBD. They concluded that all IBD patients, including patients on immune-modifying medications, should be vaccinated against COVID-19. Studies have reported that patients with IBD, including those on biologic therapy, did not have increased risk of flares post-COVID-19 immunization (31). Strong antibody responses were found in the participants who received any COVID-19 vaccine; therefore, patients with IBD are expected to develop immunity from any of the available vaccine strategies, regardless of treatment with immune-modifying therapies (32). Moreover, a position statement by the British Society of Gastroenterology section and IBD Clinical Research Group strongly supported COVID-19 vaccination for patients with IBD and stated that the risk of vaccine side effects in these patients are very low. In theory, the only risk is the suboptimal vaccine response due to treatment with immunosuppressive drugs (33). A literature review concluded that patients with IBD should be in remission prior to receiving the COVID-19 vaccine. They also recommended that their corticosteroid dose be reduced before they receive the vaccine to minimize the theoretical risk of suboptimal vaccine response (34). In two regional studies that examine the serological response of IBD patients on biologic therapies to receiving the COVID-19 vaccines, it was found that patients seroconverted after two doses of either BNT162b2 or ChAdOx1 nCoV-19 vaccines, which emphasises that despite being treated with biologic therapie, receiving the COVID-19 vaccines can be effective (35,36).

COVID-19 infection can trigger a cytokine storm syndrome, which leads to worsening of COVID-19 symptoms in the form of fevers, confusion, and coagulopathy (37). It was first hypothesized that anticytokine biologic therapies would worsen COVID-19-related outcomes; however, it was later thought that these biologic therapies which inhibit cytokine storms, could be of possible benefit (38). This is possible due to the mechanism of action of these medications. In the studies conducted around the aforementioned hypotheses, it was evident that the role of biologic therapies in COVID-19 infected patients may in fact be a protective one (5). In one of the studies conducted, it was found that patients receiving biologic therapies, in particular patients with IBD, were 5 times less likely to be diagnosed with COVID-19 infection (39). A case series from New York showed no increased risk of adverse effects in patients on biologic therapies who contract COVID-19 infection (40). This is in agreement with more studies from the literature that suggest continuation of biologic therapies, even in areas of COVID-19 outbreaks (41–43).Patients with immune-mediated diseases on biologic therapies are not at increased risk of contracting COVID-19 infection, nor are they at higher risk of severe course of the infection (44). Given the benefit of biologic therapies found in the management of auto-immune diseases, it was postulated that their use be extended to the treatment of severe COVID-19 infection or patients who develop a macrophage activation syndrome-like hyperinflammatory state or cytokine storm (45).

Since the start of the pandemic, there has been a wave of misinformation, rumors, and conspiracy theories on social media platforms, such as the perception that COVID-19 vaccines alter human DNA, that a microchip is implanted in the patient’s body when they’re injected, or that COVID-19 vaccines are live vaccines that can cause more harm than benefit (46). This has led to anti-vaccination movements worldwide. Such movements have been found to be associated with lower rates of vaccine acceptance during pandemics and disease outbreaks (47). The literature is in line with this in that study participants with IBD listed the fast development of COVID-19 vaccine, fear of worsening of their underlying disease, and concern about COVID-19 vaccine adverse effects as the main reasons they were hesitant to receive it (4,14). More reasons for COVID-19 vaccine hesitancy include: thoughts that they do not need the vaccine for protection as the nature of COVID-19 is harmless, being opposed to vaccinations in general, lack of trust in the health care system, uncertainty regarding COVID-19 vaccine efficiency, considering themselves already immunized through prior infection, and preferance to wait for other types of vaccines (48).

To see if these findings can be extended to the general patient population, larger studies in patients of diverse demographics are required. Moreover, the results of this study can be used to aid physicians (especially IBD specialists), researchers, and public health departments to develop and apply interventions to overcome COVID-19 vaccine hesitancy.

Our study is a well-designed cross-sectional study targeting patients with IBD on biologics. It evaluates factors that are associated with unwillingness to vaccinate in this vulnerable population. Also, it is the first study in Kuwait to evaluate the prevalence of vaccination among IBD patients. However, a relatively small sample size is a limitation to our study. In addition, the prevalence of vaccination in patients with IBD on other biologics (such as adalimumab and ustekinumab) were not evaluated.

## 5. Conclusion

The overall prevalence of COVID-19 vaccination among patients with inflammatory bowel disease (IBD) on biologic therapies was lower than the general population and world health organization (WHO) recommendation. Female patients, patients above the age of 50, and expatriates were more likely to be vaccinated. On the other hand, male patients, patients below the age of 50, and citizens were less likely to be vaccinated.

## Data Availability

All data produced in the present study are available upon reasonable request to the authors

## Supplementary Materials

None

## Author Contributions

**Mohammad Shehab:** Conceptualization, methodology, software, validation, formal analysis, investigation, resources, data curation, writing—original draft preparation, writing—review and editing.

**Yasmin Zurba, Ali Al Abdulsalam:** writing—original draft preparation, writing—review and editing.

**Ahmad Alfadhli, Sara Elouali:** validation, supervision, project administration.

All authors have read and agreed to the published version of the manuscript.

## Institutional Review Board Statement

The study was approved by the standing committee for coordination of health and medical research of the Ministry of health of Kuwait (IRB 2021/1729) as per the updated guidelines of the Declaration of Helsinki (64th WMA General Assembly, Fortaleza, Brazil, October 2013) and of the US Federal Policy for the Protection of Human Subjects.

## Informed Consent Statement

Patients’ informed written consent was obtained before recruitment.

## Acknowledgments

None

## References

1. Cucinotta D, Vanelli M. WHO Declares COVID-19 a Pandemic. Acta Bio Medica@: Atenei Parmensis [Internet]. 2020 [cited 2021 Nov 4];91(1):157. Available from: /pmc/articles/PMC7569573/

2. Cihan P. Forecasting fully vaccinated people against COVID-19 and examining future vaccination rate for herd immunity in the US, Asia, Europe, Africa, South America, and the World. Applied Soft Computing [Internet]. 2021 Nov 1 [cited 2021 Nov 6];111:107708. Available from: /pmc/articles/PMC8278839/

3. MacDonald NE, Eskola J, Liang X, Chaudhuri M, Dube E, Gellin B, et al. Vaccine hesitancy: Definition, scope and determinants. Vaccine. 2015 Aug 14;33(34):4161–4.

4. Costantino A, Noviello D, Conforti FS, Aloi M, Armuzzi A, Bossa F, et al. COVID-19 Vaccination Willingness and Hesitancy in Patients With Inflammatory Bowel Diseases: Analysis of Determinants in a National Survey of the Italian IBD Patients’ Association. Inflammatory Bowel Diseases [Internet]. 2021 Jul 14 [cited 2021 Nov 6];1–5. Available from: https://academic.oup.com/ibdjournal/advance-article/doi/10.1093/ibd/izab172/6321213

5. Alrashed F, Battat R, Abdullah I, Charabaty A, Shehab M. Impact of medical therapies for inflammatory bowel disease on the severity of COVID-19: a systematic review and meta-analysis. BMJ Open Gastroenterology [Internet]. 2021 Oct 1 [cited 2021 Nov 6];8(1):e000774. Available from: https://bmjopengastro.bmj.com/content/8/1/e000774

6. Wong S-Y, Dixon R, Pazos VM, Gnjatic S, Colombel J-F, Cadwell K, et al. Serologic Response to Messenger RNA Coronavirus Disease 2019 Vaccines in Inflammatory Bowel Disease Patients Receiving Biologic Therapies. Gastroenterology [Internet]. 2021 Aug 1 [cited 2021 Nov 6];161(2):715. Available from: /pmc/articles/PMC8055494/

7. Kennedy NA, Lin S, Goodhand JR, Chanchlani N, Hamilton B, Bewshea C, et al. Infliximab is associated with attenuated immunogenicity to BNT162b2 and ChAdOx1 nCoV-19 SARS-CoV-2 vaccines in patients with IBD. Gut [Internet]. 2021 Oct 1 [cited 2021 Nov 6];70(10):1884–93. Available from: https://gut.bmj.com/content/70/10/1884

8. COVID-19 and IBD Reporting Database | SECURE-IBD Database [Internet]. [cited 2021 Nov 29]. Available from: https://covidibd.org/

9. Brenner EJ, Ungaro RC, Gearry RB, Kaplan GG, Kissous-Hunt M, Lewis JD, et al. Corticosteroids, But Not TNF Antagonists, Are Associated With Adverse COVID-19 Outcomes in Patients With Inflammatory Bowel Diseases: Results From an International Registry. Gastroenterology [Internet]. 2020 Aug 1 [cited 2021 Nov 29];159(2):481. Available from: /pmc/articles/PMC7233252/

10. Rubin DT, Abreu MT, Rai V, Siegel CA, Disease IO for the S of IB. Management of Patients With Crohn’s Disease and Ulcerative Colitis During the Coronavirus Disease-2019 Pandemic: Results of an International Meeting. Gastroenterology [Internet]. 2020 Jul 1 [cited 2021 Nov 6];159(1):6. Available from: /pmc/articles/PMC7194599/

11. covidvax.live - Kuwait [Internet]. [cited 2021 Nov 26]. Available from: https://covidvax.live/location/kwt

12. Comirnaty and Pfizer-BioNTech COVID-19 Vaccine | FDA [Internet]. [cited 2021 Nov 8]. Available from: https://www.fda.gov/emergency-preparedness-and-response/coronavirus-disease-2019-covid-19/comirnaty-and-pfizer-biontech-covid-19-vaccine#additional

13. ICD-10 Version:2019 [Internet]. [cited 2021 Nov 8]. Available from: https://icd.who.int/browse10/2019/en#/K52

14. Vieira Rezende RP, Braz AS, Guimarães MFB, Ribeiro SLE, Abreu Vieira RMR, Bica BE, et al. Characteristics associated with COVID-19 vaccine hesitancy: A nationwide survey of 1000 patients with immune-mediated inflammatory diseases. Vaccine. 2021 Oct 22;39(44):6454–9.

15. COVID-19: Science in 5: Episode #1 - Herd immunity [Internet]. [cited 2021 Nov 26]. Available from: https://www.who.int/emergencies/diseases/novel-coronavirus-2019/media-resources/science-in-5/episode-1

16. Wang W, Wu Q, Yang J, Dong K, Chen X, Bai X, et al. Global, regional, and national estimates of target population sizes for covid-19 vaccination: descriptive study. BMJ [Internet]. 2020 Dec 15 [cited 2021 Nov 26];371. Available from: https://www.bmj.com/content/371/bmj.m4704

17. Alaskar D, AlAmeel T, al Besher M, al Sulais E. Attitude of patients with IBD toward COVID-19 vaccine in Saudi Arabia. United European Gastroenterology Journal [Internet]. 2021 Oct [cited 2021 Nov 26];9(S8):530–530. Available from: https://doi.org/10.1002/ueg2.12144

18. Bertakis KD, Azari R, Helms LJ, Callahan EJ, Robbins JA. Gender differences in the utilization of health care services. The Journal of family practice [Internet]. 2000 [cited 2021 Nov 30];49(2):147–52. Available from: https://pubmed.ncbi.nlm.nih.gov/10718692/

19. Vassallo A, Shajahan S, Harris K, Hallam L, Hockham C, Womersley K, et al. Sex and Gender in COVID-19 Vaccine Research: Substantial Evidence Gaps Remain. Frontiers in Global Women’s Health [Internet]. 2021 Nov 1 [cited 2021 Nov 30];2. Available from: /pmc/articles/PMC8593988/

20. Priori R, Pellegrino G, Colafrancesco S, Alessandri C, Ceccarelli F, di Franco M, et al. SARS-CoV-2 vaccine hesitancy among patients with rheumatic and musculoskeletal diseases: a message for rheumatologists. Annals of the Rheumatic Diseases [Internet]. 2021 Jul 1 [cited 2021 Nov 10];80(7):953–4. Available from: https://ard.bmj.com/content/80/7/953

21. Selim R, Wellens J, Marlow L, Satsangi JJ. SARS-CoV-2 vaccination uptake by patients with inflammatory bowel disease on biological therapy. The Lancet Gastroenterology & Hepatology [Internet]. 2021 Oct [cited 2021 Nov 10];0(0). Available from: http://www.thelancet.com/article/S2468125321003472/fulltext

22. Walldorf J, Arnim U von, Schmelz R, Riesner-Wehner A, Michl P, Grunert PC, et al. SARS-CoV-2 Vaccination in Patients With Inflammatory Bowel Disease—Fear and Desire. Inflammatory Bowel Diseases [Internet]. 2021 Jun 28 [cited 2021 Nov 30];1–4. Available from: /pmc/articles/PMC8344527/

23. COVID-19 vaccines, pregnancy and breastfeeding [Internet]. [cited 2021 Nov 11]. Available from: https://www.rcog.org.uk/en/guidelines-research-services/coronavirus-covid-19-pregnancy-and-womens-health/covid-19-vaccines-and-pregnancy/covid-19-vaccines-pregnancy-and-breastfeeding/

24. COVID-19 Vaccination Considerations for Obstetric–Gynecologic Care | ACOG [Internet]. [cited 2021 Nov 11]. Available from: https://www.acog.org/clinical/clinical-guidance/practice-advisory/articles/2020/12/covid-19-vaccination-considerations-for-obstetric-gynecologic-care

25. Blakeway H, Prasad S, Kalafat E, Heath PT, Ladhani SN, le Doare K, et al. COVID-19 vaccination during pregnancy: coverage and safety. American Journal of Obstetrics and Gynecology [Internet]. 2021 [cited 2021 Nov 30]; Available from: /pmc/articles/PMC8352848/

26. Helen Skirrow A, Barnett S, Bell S, Riaposova L, Mounier-Jack S, Kampmann B, et al. Women’s views on accepting COVID-19 vaccination during and after pregnancy, and for their babies: A multi-methods study in the UK. medRxiv [Internet]. 2021 May 3 [cited 2021 Nov 30];2021.04.30.21256240. Available from: https://www.medrxiv.org/content/10.1101/2021.04.30.21256240v1

27. Adhikari EH, Spong CY. COVID-19 Vaccination in Pregnant and Lactating Women. JAMA [Internet]. 2021 Mar 16 [cited 2021 Nov 30];325(11):1039–40. Available from: https://jamanetwork.com/journals/jama/fullarticle/2776449

28. Martins I, Louwen F, Ayres-de-Campos D, Mahmood T. EBCOG position statement on COVID-19 vaccination for pregnant and breastfeeding women. European Journal of Obstetrics, Gynecology, and Reproductive Biology [Internet]. 2021 Jul 1 [cited 2021 Nov 30];262:256. Available from: /pmc/articles/PMC8120807/

29. Lee YJ, Kim SE, Park YE, Chang JY, Song HJ, Kim DH, et al. SARS-CoV-2 Vaccination for Adult Patients with Inflammatory Bowel Disease: Expert Consensus Statements by KASID. The Korean Journal of Gastroenterology [Internet]. 2021 Aug 25 [cited 2021 Nov 30];78(2):117–28. Available from: https://www.kjg.or.kr/journal/view.html?doi=10.4166/kjg.2021.110

30. International Organization For the Study of Inflammatory Bowel Disease | To promote the health of people with IBD worldwide [Internet]. [cited 2021 Nov 30]. Available from: https://ioibd.org/

31. Botwin GJ, Li D, Figueiredo J, Cheng S, Braun J, McGovern DPB, et al. Adverse Events After SARS-CoV-2 mRNA Vaccination Among Patients With Inflammatory Bowel Disease. The American journal of gastroenterology [Internet]. 2021 Aug [cited 2021 Nov 30];116(8):1746–51. Available from: https://pubmed.ncbi.nlm.nih.gov/34047304/

32. Siegel CA, Melmed GY, McGovern DPB, Rai V, Krammer F, Rubin DT, et al. SARS-CoV-2 vaccination for patients with inflammatory bowel diseases: recommendations from an international consensus meeting. Gut [Internet]. 2021 Apr 1 [cited 2021 Nov 30];70(4):635. Available from: /pmc/articles/PMC7818789/

33. Alexander JL, Moran GW, Gaya DR, Raine T, Hart A, Kennedy NA, et al. SARS-CoV-2 vaccination for patients with inflammatory bowel disease: a British Society of Gastroenterology Inflammatory Bowel Disease section and IBD Clinical Research Group position statement. The Lancet Gastroenterology and Hepatology [Internet]. 2021 Mar 1 [cited 2021 Nov 30];6(3):218–24. Available from: http://www.thelancet.com/article/S2468125321000248/fulltext

34. Doherty J, Fennessy S, Stack R, O’ Morain N, Cullen G, Ryan EJ, et al. Review Article: vaccination for patients with inflammatory bowel disease during the COVID-19 pandemic. Alimentary pharmacology & therapeutics [Internet]. 2021 Nov 1 [cited 2021 Nov 30];54(9):1110–23. Available from: https://pubmed.ncbi.nlm.nih.gov/34472643/

35. Shehab M, Abu-Farha M, Alrashed F, Alfadhli A, Alotaibi K, Alsahli A, et al. Immunogenicity of BNT162b2 Vaccine in Patients with Inflammatory Bowel Disease on Infliximab Combination Therapy: A Multicenter Prospective Study. Journal of clinical medicine [Internet]. 2021 Nov 18 [cited 2021 Dec 1];10(22):5362. Available from: https://pubmed.ncbi.nlm.nih.gov/34830644/

36. Shehab M, Alrashed F, Alfadhli A, Alotaibi K, Alsahli A, Mohammad H, et al. Serological Response to BNT162b2 and ChAdOx1 nCoV-19 Vaccines in Patients with Inflammatory Bowel Disease on Biologic Therapies; A Multi-Center Prospective Study. medRxiv [Internet]. 2021 Nov 1 [cited 2021 Dec 1];2021.10.31.21265718. Available from: https://www.medrxiv.org/content/10.1101/2021.10.31.21265718v1

37. Wang D, Hu B, Hu C, Zhu F, Liu X, Zhang J, et al. Clinical Characteristics of 138 Hospitalized Patients With 2019 Novel Coronavirus–Infected Pneumonia in Wuhan, China. JAMA [Internet]. 2020 Mar 17 [cited 2021 Nov 30];323(11):1061. Available from: /pmc/articles/PMC7042881/

38. Yousaf A, Gayam S, Feldman S, Zinn Z, Kolodney M. Clinical outcomes of COVID-19 in patients taking tumor necrosis factor inhibitors or methotrexate: A multicenter research network study. Journal of the American Academy of Dermatology [Internet]. 2021 Jan 1 [cited 2021 Nov 30];84(1):70–5. Available from: http://www.jaad.org/article/S0190962220325937/fulltext

39. Bezzio C, Pellegrini L, Manes G, Arena I, Picascia D, Corte C della, et al. Biologic Therapies May Reduce the Risk of COVID-19 in Patients With Inflammatory Bowel Disease. Inflammatory Bowel Diseases [Internet]. 2020 Oct 1 [cited 2021 Nov 30];26(10):E107–9. Available from: /pmc/articles/PMC7499633/

40. Haberman R, Axelrad J, Chen A, Castillo R, Yan D, Izmirly P, et al. Covid-19 in Immune-Mediated Inflammatory Diseases — Case Series from New York. New England Journal of Medicine [Internet]. 2020 Jul 2 [cited 2021 Nov 30];383(1):85–8. Available from: https://www.nejm.org/doi/10.1056/NEJMc2009567

41. Megna M, Napolitano M, Patruno C, Fabbrocini G. Biologics for psoriasis in COVID-19 era: What do we know? Dermatologic Therapy [Internet]. 2020 Jul 1 [cited 2021 Nov 30];33(4):e13467. Available from: https://onlinelibrary.wiley.com/doi/full/10.1111/dth.13467

42. Bashyam AM, Feldman SR. Should patients stop their biologic treatment during the COVID-19 pandemic. The Journal of dermatological treatment [Internet]. 2020 May 18 [cited 2021 Nov 30];31(4):317–8. Available from: https://pubmed.ncbi.nlm.nih.gov/32191143/

43. Magnano M, Balestri R, Bardazzi F, Mazzatenta C, Girardelli CR, Rech G. Psoriasis, COVID-19, and acute respiratory distress syndrome: Focusing on the risk of concomitant biological treatment. Dermatologic therapy [Internet]. 2020 Jul 1 [cited 2021 Nov 30];33(4). Available from: https://pubmed.ncbi.nlm.nih.gov/32475056/

44. Brazzelli V, Isoletta E, Barak O, Barruscotti S, Vassallo C, Giorgini C, et al. Does therapy with biological drugs influence COVID-19 infection? Observational monocentric prevalence study on the clinical and epidemiological data of psoriatic patients treated with biological drugs or with topical drugs alone. Dermatologic Therapy [Internet]. 2020 Nov 1 [cited 2021 Nov 30];33(6):e14516. Available from: https://onlinelibrary.wiley.com/doi/full/10.1111/dth.14516

45. González-Gay MA, Castañeda S, Ancochea J. Biologic Therapy in COVID-19. Archivos De Bronconeumologia [Internet]. 2021 Jan 1 [cited 2021 Nov 30];57:1. Available from: /pmc/articles/PMC7318980/

46. Biswas MR, Alzubaidi S, Shah U, Abd-Alrazaq AA, Shah Z, Barattucci M. A Scoping Review to Find Out Worldwide COVID-19 Vaccine Hesitancy and Its Underlying Determinants. Vaccines 2021, Vol 9, Page 1243 [Internet]. 2021 Oct 25 [cited 2021 Nov 11];9(11):1243. Available from: https://www.mdpi.com/2076-393X/9/11/1243/htm

47. Dubé E, Vivion M, MacDonald NE. Vaccine hesitancy, vaccine refusal and the anti-vaccine movement: influence, impact and implications. http://dx.doi.org/101586/147605842015964212 [Internet]. 2014 Jan 1 [cited 2021 Nov 11];14(1):99–117. Available from: https://www.tandfonline.com/doi/abs/10.1586/14760584.2015.964212

48. Troiano G, Nardi A. Vaccine hesitancy in the era of COVID-19. Public health [Internet]. 2021 May 1 [cited 2021 Nov 30];194:245–51. Available from: https://pubmed.ncbi.nlm.nih.gov/33965796/

